# Gender Disparity in Occupational Health Hazards and Risks Among Coastal Fisherfolk in Chattogram and Cox’s Bazar Districts in Bangladesh: A comparative mix-method cross-sectional study

**DOI:** 10.1101/2025.07.01.25330377

**Authors:** Charls Erik Halder, Md Abeed Hasan, Anannya Roy, Liton Chandra Bhoumick, Rajib Das, Sourav Nath Mithun, S.M. Tareq Rahman, Hamim Tassdik, Partha Pratim Das, Mohammad Delwer Hossain Hawlader

**Affiliations:** Public Health and Research Division, Bright Bangladesh Forum, Chattogram, Bangladesh; UFR Life Science, Universite Paris Cite, Paris, France; Department of Public Health, North South University, Dhaka, Bangladesh; Department of Public Health, University of South Asia, Dhaka, Bangladesh; Faculty of Medicine and Life Sciences, University of Chester, Chester, UK; Department of Public Health, American International University-Bangladesh (AIUB), Dhaka, Bangladesh; Department of Public Health, State University of Bangladesh, Dhaka, Bangladesh; Public Health and Research Division, Anvesion, Dhaka, Bangladesh

**Keywords:** Gender disparities, Occupational health, Coastal fisheries, Bangladesh, Fisherfolk, Workplace safety, Public health equity

## Abstract

**Background:** In coastal Bangladesh, while men predominantly engage in direct fishing from rivers and the Bay of Bengal, women are mostly engaged in fish processing. Due to the different roles of men and women in fisheries, occupational hazards and risks can also be distinctive. However, there is limited comparative evidence on the role of gender in occupational hazards and risks among men and women working in fisheries sector.

**Objective:** This study investigated and compared the prevalence, type, and severity of occupational health hazards and associated risks among male and female fisherfolk in Chattogram and Cox’s Bazar, Bangladesh.

**Methods:** An explanatory sequential mixed-methods design was used. The qualitative component included nine focus group discussions on gendered exposures and hazards, which informed a subsequent cross-sectional survey of 407 fisherfolk (n = 200 males; n = 207 females). Gender-disaggregated responses were analysed using descriptive statistics, chi-square tests, and radar charts. Key qualitative themes were developed and used for cross-method triangulation.

**Results:** In the fish industry, male workers were found with a high proportion of engagement in direct fishing (male 52.5% vs. female 0%, p < 0.001), whereas female workers were majorly found involved in dry fish processing (54.1% vs. 14.5%, p < 0.001). Most risk factors such as sun and salt exposure and lack of sanitary latrines—were reported by both men and women at consistently high rates. Females were found having higher chemical exposure (53.6% vs. 35.0%; p < 0.001) and males had higher exposure to heavy weight (93.0% vs. 82.6%, p = 0.002). A high prevalence of occupational risks including skin diseases, musculoskeletal issues, diarrhoea, reported among both groups with comparable risks. Conditions that were significantly higher among male workers were fish stings or bites, motion sickness and earache. Inaccessibility to toilets, absence of menstrual hygiene kits, lack of breastfeeding space and lack of maternity leaves were some of the unique concerns of women. Drowning and falling overboard, cyclones and storms and attacks from pirates are unique hazards for men. Men are more prone to fatality due to their engagement in acute life-threatening tasks. The use of PPE is low among both men and women, with 65% to 99% reporting not using the specific PPEs.

**Conclusions:** This study revealed a high burden of occupational hazards and risks among both male and female workers in the fisheries sector in Bangladesh from a gender lens. A gender-sensitive occupational safety and health strategy and action plan strategy is crucial to attain the health, wellbeing, dignity, productivity, and protection of the men and women in fisherfolk communities in Bangladesh.

## INTRODUCTION

In 2022, marine fisheries and aquaculture collectively produced approximately 223 million tons of aquatic animals, highlighting their critical role in the provision of food security and nutrition globally (1). An estimated 61.8 million people globally rely predominantly on fisheries, aquaculture, and associated activities for their livelihoods, although the number is somewhat greater if we consider other associated activities, such as processing (2). In Bangladesh, approximately 17 million people rely on fishing, aquaculture, fish handling, and processing (3). Regardless of the significant contributions of fishing to food security, nutrition, and the economy, it remains one of the most hazardous industries, resulting in thousands of injuries, diseases, and fatalities.

The characteristics of employment and nature of works in fisheries are highly gendered. Men and women are engaged in diverse areas of fisheries and aquaculture, which widely vary from region to region (4). In coastal Bangladesh, while men predominantly engage in direct fishing from rivers and the Bay of Bengal, women are mostly engaged in fish sorting, grading, cutting, dry fish processing, dry fish paste (Nappi) processing, daily labouring in fish farms, transporting, selling fish or fish products, net making, ice processing, fish basket making, and collecting minnows and seashells from the sea shores (5). Due to the different roles of men and women in fisheries, occupational hazards and risks are also distinctive.

Gender refers not to male and female, but rather to the “social attributes and opportunities associated with being male and female” (6). Women in fishing communities often carry a dual burden, balancing income-generating roles in fisheries with traditional household and caregiving responsibilities (5). Despite heavy workload, the role of women in fisheries is often not recognized. On the other hand, societal and cultural expectations from men lead them to engage in risk-taking and life-threatening activities as affirmation of their masculine identity (7). For instance, in Bangladesh, around 255 trawlers and 67,699 mechanized and non-mechanized boats are engaged in fishing only in marine fisheries (8). In the coastal areas of the country, sinking, capsizing, disappearing, and floating-away boats are common occurrences that frequently result in the deaths or disappearances of fishermen (9). Long working hours, extreme weather, sinking, capsizing, operation of unfitted or inappropriately geared vessels, and handling of heavy machinery contribute to high mortality of workers, especially fishermen, due to their engagement in direct fishing (8,9). Therefore, the differences in social attributes, opportunities, differences, and expectations may also have an influence on the occupational health and well-being of the workers in the fisheries.

Rahman et al. (2024) using the Pro-WEFI index, found only 14% of women in Bangladesh’s coastal fishing communities were empowered, compared to 37% of men (10). The Gender Parity Index (GPI) stood at 0.79, with only 31% of households having parity. The greatest gaps were in decision-making, income control, and mobility, directly impacting women’s ability to seek health services or negotiate for safer working conditions. It further stressed that gender inequality in access to education, capital, and land rights increases occupational vulnerability among coastal women (11). This suggests that occupational health is not just technical but deeply social-mediated by power, roles, and cultural expectations. There could be some hidden hazards which remain under-reported. Mangubhai et al. (2023) argued that gender-based violence and sexual harassment, economic coercion, and emotional abuse are occupational hazards in fisheries (12). Their global review noted that such violence is reinforced by institutional blindness, tolerated due to cultural norms, and unaddressed in fisheries governance.

In our first article of an umbrella research initiative on occupational hazards among the coastal fishing community, we presented the high prevalence of occupational hazards and risks among the women in fisherfolk. (13) This include physical safety hazards such as slippery surfaces and fish-cutting instruments; physical hazards like prolonged sun exposure and noise; chemical hazards like pesticides and saltwater; ergonomic hazards such as prolonged uncomfortable posture and heavy lifting; and biological hazards including inadequate sanitation facilities (13). We also identified risks resulting from the hazards, including injuries, musculoskeletal conditions, skin diseases/conditions, eye complaints, severe respiratory distress, and a high incidence of self-reported communicable diseases (13). Several studies in similar settings in developing countries, like India and Kenya, also found similar occupational hazards and risks among women working in the fisheries sector (14–16). Several studies reported hazards among fishermen, which include the possibility of wrecking or capsizing the fishing boats, fires or explosions at the board or workplace, falling due to slippery surfaces, entanglement in nets or other gear, and unsafe handling of heavy, dangerous, or unguarded equipment (12,17). Exposure to sun, prolonged physical labour, exposure to saline water, and occupational accidents are among the reported hazards, and red eye conditions, musculoskeletal issues, and waterborne infections are commonly reported risks among fishermen (18). While both men and women may share some similar hazards and risks, due to their different roles and level of engagement, the type, severity, and likelihood of the hazards and risks could significantly vary.

However, there is limited comparative evidence on the role of gender in the fisheries sector, with only a few studies that directly compared the hazards and risks between male and female workers within the same fishing communities. A few studies found that fishermen without personal protective equipment (PPE) are significantly more likely to experience hazards (19). Our previous study reported that most women (78.26%) in the fishing community do not use personal protective equipment, and the majority of them (93.72%) lack a workplace first aid kit. However, there are limited studies available regarding the availability and use of PPE among the male counterparts in fishing communities in coastal Bangladesh.

Understanding how occupational hazards, risks, and protective measures vary by gender is crucial for designing effective health and safety interventions for the marginalized men and women workers in the fisheries sector. Such comparative analysis can inform gender-sensitive policies and promote equitable access to occupational health resources in fishing communities. Therefore, this study has been undertaken under the same umbrella research initiative of our previous article, expanding the exploration of the hazards and risks among fishermen from a gender-comparative perspective. The overall objective of this study was to compare the prevalence of occupational risk factors and hazards among male and female workers in the fisheries sector in Chattogram and Cox’s Bazar, Bangladesh. While we used the same dataset previously used to the study of women in fishing community, comparable data have been gathered from fishermen to generate a comparative analysis from a gender lens, highlighting the gender-specific disparities and commonalities in occupational exposures.

## METHODOLOGY

Our study adopted an explanatory sequential mixed-method design to assess gendered occupational health hazards among coastal fisherfolk in Bangladesh. This is an extension of our earlier research that focused exclusively on fisherwomen. In this study, we added data from men in the fishing community to support a comparative gender perspective. The study was conducted in Chattogram and Cox’s Bazar, the two districts in Bangladesh with extensive fishing activities.

### Study Sites

Thousands of fisherfolk communities reside along the Bay of Bengal, and our selected sites in Cox’s Bazar and Chattogram represent a cross-section of the fishing sector in Bangladesh. In Cox’s Bazar, fishing at sea and coastal fish processing are the primary sources of income. In Chattogram, fishing activity is concentrated along the Karnaphuli River where large colonies of fisherfolk are involved in both catching and processing. Furthermore, these two districts were the locations of our previously conducted research, so they serve as a valid comparison over time across gender. A more detailed socio-demographic situation and map of the study location is available here (see Fig 1) (13). A total 304 participants individuals from Cox’s Bazar and 103 individuals from Chattogram, who primarily dependent on fisheries activities participated in this study (Table 1).

**Figure 1.** Map of study locations in Cox’s Bazar and Chattogram. Geographical distribution of selected fisherfolk communities included in the study, showing eight coastal sites.

**Table 1:**
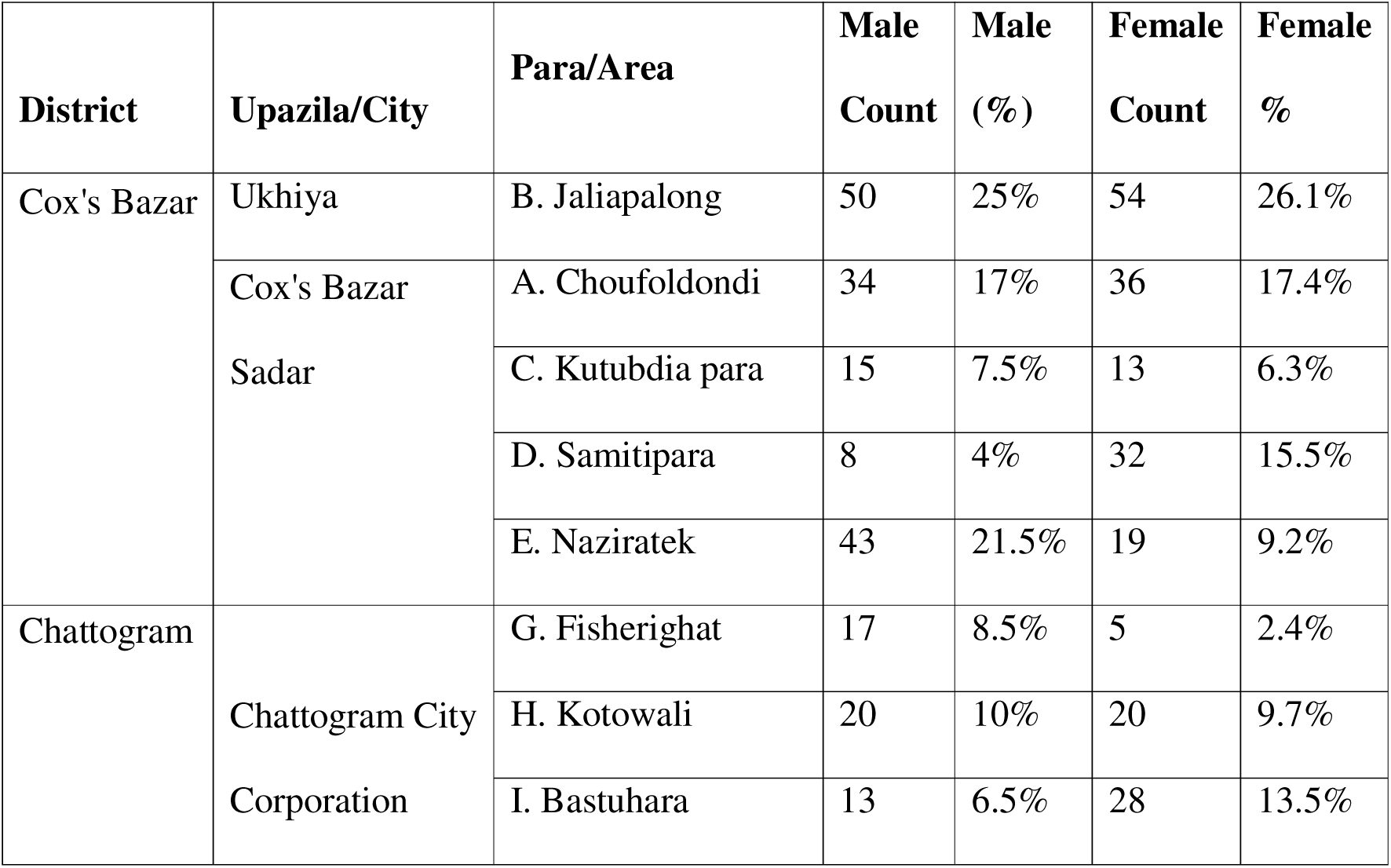
Distribution of Study Participants by District, Upazila, and Village/Area Among Coastal Fisherfolk in Cox’s Bazar and Chattogram, Bangladesh

### Study design, sampling and data collection

This is a cross-sectional study with *explanatory sequential mixed methods* design, where quantitative data collection was conducted first to identify a quantitative aspect of occupational hazards, risks and the status of safety measures among men and women, followed by a qualitative data collection was carried out to further explain the quantitative findings (20). The data was conducted in iterative rounds between September 2023 and February 2024 to explore contextual factors for the results found.

Followed by the data collection, the integration of results from both methods were conducted in a structured sequence to get a deeper insight into both results. The integrative approach enhances understanding of how gender shapes exposure, impact, and coping mechanisms within fisheries work, consistent with best practices in mixed-method research (Fetters et al., 2013).

In the quantitative component, we surveyed 407 participants (200 males and 207 females) using a structured questionnaire. The study was implemented in the same eight selected sites in Cox’s Bazar and Chattogram we covered in the last study, where approximately 7,000 fisherfolk families are residing (13). Similar to the previous study for the women, sample size for male participants was calculated using an 85% occupational injury rate from a similar setting as a reference point (21). Applying Cochran’s formula, the sample size required for a 95% confidence level and a 5% sampling error was 191 for men. However, we have adjusted the final sample size to 200 participants considering the potentiality of unresponsiveness. We have selected the participants using simple random sampling from a local list of fisher families.

In the quantitative component of the study, we conducted a survey of women (previous study) and men (extension of the previous study) working in the fisheries sector using a structured questionnaire (13). We deployed the questionnaire through the Kobo toolbox for data collection. The questionnaire included questions in relation to socio-demographic profile of the participants, exposure to different types of occupational hazards, potential risks of the occupational hazards and status of different health and safety measures. A detailed of the questionnaire for female workers is available here (13), while for the men it is annexed as S1 file. The questionnaires were initially piloted respectively among 10 men and 10 women, and necessary adjustments were carried out to finalize the questionnaire. The same four trained enumerators who worked in the previous study collected the data. They were fluent in the local Chittagongian language and worked under the close supervision of the research assistants.

For the qualitative component of the study, we have collected the data through nine focus group discussions (FGDs) gendered and mixed male/female of fishing community to better understand the nature of work, hazards and risks of their occupation, and gender dimensions of their occupational hazards and risks. A total of 48 men and 55 women from the fisherfolk communities in Chattogram and Cox’s Bazar participated in the focus group discussions.

Each of the FGD were participated in by six to twelve participants. With the assistance of our volunteers and local stakeholders in fisheries, the participants were purposively selected based on their engagement in different fishing roles. Table 2 demonstrates the demographic characteristics of the FGD participants.

**Table 2:** Demographics of the participants in FGD

| Type of industry | Male |  |  |  | Female |  |  |  |
| --- | --- | --- | --- | --- | --- | --- | --- | --- |
|  | <18<br>years | 18 –<br>30<br>years | 31 –<br>60<br>years | >60<br>years | <18<br>years | 18 –<br>30<br>years | 31 –<br>60<br>years | >60<br>years |
| Direct fishing | 0 | 5 | 8 | 1 | 0 | 1 | 2 | 0 |
| Dry fish processing | 0 | 3 | 4 | 0 | 1 | 5 | 6 | 2 |
| Powdered Dry Fish<br>(Nappi) processing | 0 | 2 | 2 | 0 | 0 | 4 | 3 | 0 |
| Raw fish processing/<br>marketing | 0 | 3 | 5 | 0 | 0 | 3 | 5 | 1 |

The semi-structured guiding questionnaire (see S2 file) in the previous study was adapted to cover the themes of occupational activities, hazards, risks, safety measures and gender-specific concerns among both men and women in the fisher communities. The questionnaire also included questions to explain the results that initially came from the quantitative surveys. The principal investigator conduced the FGDs and 5 trained research assistants. The research assistants were distributed with the role of consent taking, note-taking and audio recording, local coordination, maintaining group dynamics, and observing non-verbal cues. The research team engaged interpreters fluent in the local Chittagongian language to establish effective communication. Duration of the FGDs ranged from 90 to 120 minutes.

### Data Analysis

Following the cleaning of the collected quantitative data, only frequently reported risks were retained; rare, inconsistent, or outlier responses were excluded to improve analytical clarity. Descriptive statistics were used to summarise the most common hazards, and radar charts visualized gender-specific exposure patterns. Chi-square tests were conducted to assess statistically significant differences between male and female groups. The qualitative data (FGD transcripts) were transcribed verbatim. A thematic analysis of the qualitative data was conducted manually, revealing consistent, detailed and explanatory information aligned to the quantitative data, including widespread physical safety hazards, chemical exposure, ergonomic hazards, workplace injuries and fatalities, and socio-cultural challenges like the issues specific to femininity and masculinity, as well as inadequate safety protocols. This study adopts an integrative design, combining qualitative and quantitative findings to produce a comprehensive understanding. While our earlier research followed a qualitative-to quantitative sequence, this study reverses the flow moving from quantitative findings to qualitative measurement enabling detailed explanation of the gender dimension in occupational health hazards, risks and safety.

## ETHICAL CONSIDERATION

All ethical guidelines were strictly followed throughout the research process. The Institutional Review Board (IRB) and Ethics Review Committee (ERC) of North South University in Bangladesh approved the research protocol (2023/OR-NSU/IRB/0810). All participants provided written informed consent, commonly using thumbprints due to literacy barriers. Consent forms were securely stored. Our study was designed to follow the principle of “do no harm” and posed no legal risk to study participants. The study adhered strictly to the ethical principles outlined in the Helsinki Declaration (1964) and its 2013 revision.

## RESULTS

### Socio-demographic profile of the study participants

A total of 207 women and 200 men working in the fisheries sector in Cox’s Bazar and Chattogram participated in this study. Table 3 summarizes the socio-demographic profile of the participants. The majority of participants for both genders belonged to the age group 31 45 years (46.5% for male and 58% for female). Figure 2, Histogram of age distribution among men and women, demonstrates a bell-shaped age distribution in both men and women, resembling a normal distribution.

**Table 3.**
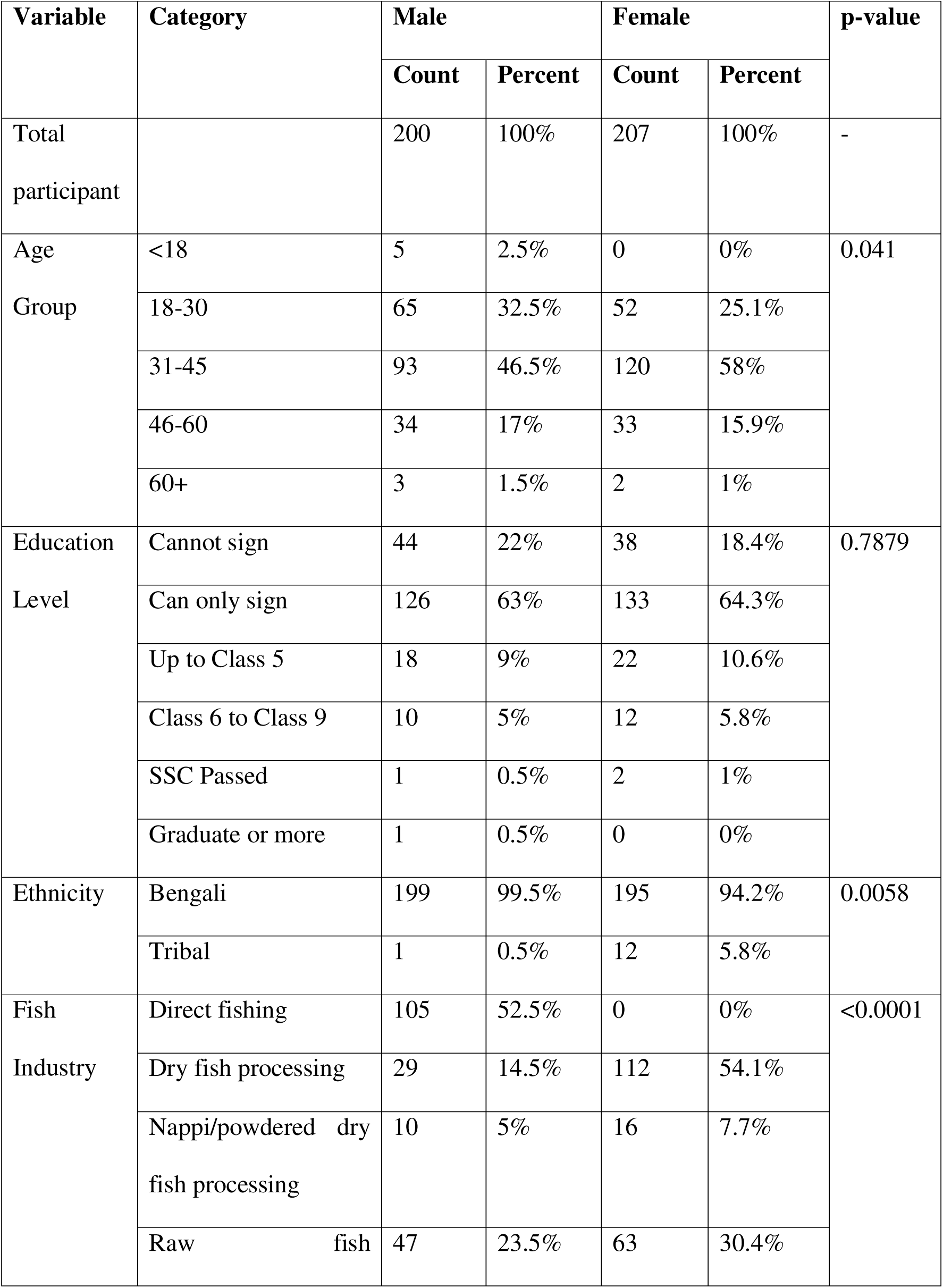
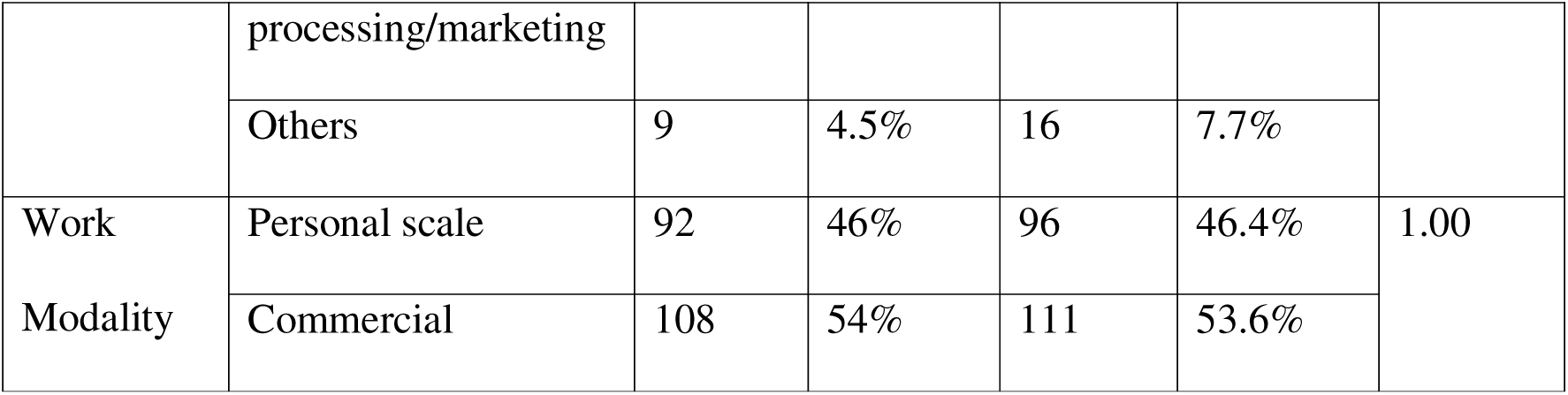
Sociodemographic Table

**Figure 2.** Age distribution histogram among men and women workers in the fishing community. Histogram showing a bell-shaped age distribution for both male and female participants.

Majority of the participants either cannot sign (male 22%, female 18.4%) or can only sign (male 63%, female 64.3%), with comparable distribution across both sexes. Most of the participants in both groups belonged to the mainstream Bengali community, whereas a small fraction was tribal, belonging to the Rakhaine community. However, within the tribal ethnic group, *Rakhaine*, the proportion of female participants was significantly higher than male (male 0.5%, female 5.8%, p = 0.0058). In the fish industry, male workers were found with an exclusively high proportion of engagement in direct fishing (male 52.5% vs. female 0%, p < 0.001), whereas female workers were majorly found involved in dry fish processing (54.1% vs. 14.5%, p < 0.001). However, both groups have their participation in other fisheries-related activities, including powdered dry fish (*Nappi*) processing and raw fish processing/marketing. Both males and females were found working both at personal scale works as well as commercial works, which did not differ significantly by sex (p = 1.00).

A gendered division of roles in fisheries was also revealed during the focus group discussions. Men in the fisherfolk community are mostly engaged in activities related to direct fishing, such as deep-sea or riverine fishing, boat operation, engine operation, and handling of nets. The work of men is considered very risky due to their extensive engagement in sea-based activities. In contrast, women mainly engaged in coast-based and processing tasks, which are often unstructured and require prolonged engagement. Common examples of their work are sorting and grading, cutting and tying, sun-drying, Nappi processing (segregating shrimps, fermenting, grinding, and drying), minnow collection, and fish selling. Women often need to carry their children to the shore and continue working till night, depending on the arrival of the boats from the sea or river. A woman shared her experience, *“We collect larvae, catch fish, wait for our husband’s boat to arrive, then we collect fish from the net, dry them, and sell them.”* This is supplemented by another woman, *“Even if we are doing household work, once the boat comes, we go quickly to collect the shrimp and other fishes” sometimes the whole night.”* Women reported that they need to carry out these economic activities in addition to maintaining their household responsibilities and caregiving roles.

There is a strong social norm where men perform the more dangerous aspects of fishing, while women are assigned to processing fish and preparing them for sale. Women experienced limited prospects for mobility within the occupation, as they were mostly restricted to the processing aspect of the industry.

There are several methods of payment for the men. For small boats, fishermen are paid on a revenue-sharing model, where payment is considered a share of the total fish caught. As expressed by one fisherman, “*It’s 50% for the boat owner and 50% for the fishermen. Among six people on board, everyone gets one share. The Majhi gets 1.5 shares as a bonus.*” However, in coast-based work, like fish processing, loading or transporting, men are paid on a daily labour basis, ranging from BDT 500 to1000. For the bigger boats, sometimes the fishermen are hired on an annual basis, where a block payment is fixed for the whole year’s engagement of the hired fishermen. In contrast, women are paid based on daily wages, ranging between BDT 200 to 400 per day, which is much lower than that of men. In many cases, payment is not given in cash; rather, they are allowed to keep a portion of the fish. In Nappi processing, women receive compensation in the form of retained medium-sized shrimp, which they subsequently sell in the local market.

Payment is usually strictly conditional for women on work attendance: *“No work, no pay is the rule here. We take leave according to our health.”* Employers do not make any payments for days missed due to sickness. There is no provision for maternity leave or provision of leaves for medical emergencies. Both men and women are hired informally, and there are no fixed contracts and no job security. Employers often do not contribute to healthcare or benefits.

Every year, there is an off-season, when the government Bans sea fishing (e.g., 65 days). Fishing is also limited during stormy weather. Usually, fishermen take a rest during this off-season. The same happens for women. They remain engaged in household chores and childcare. There is usually no income from fishing or related activities. Therefore, some men and women are found to be engaged in other activities, like larvae collection, seashell collection, and day labour. Some women reported that they engage in informal domestic housekeeping jobs.

### Occupational Hazards and Risks

Table 4 demonstrates the comparative prevalence of occupational hazards among men and women engaged in fisheries. We found a very high prevalence of sun exposure (more than 98.5%) and saltwater exposure among both male and female groups, and these differences were not statistically significant, meaning both groups are equally vulnerable to the hazards. However, females were found to be more vulnerable to chemical exposure as compared to males (53.6% vs. 35.0%). The logistic regression found a statistically significant more than twice higher odds of chemical exposure among females (OR = 2.15, 95% CI: 1.44–3.21, *p*< 0.001). Although the prevalence of the hazard of lifting heavy weight is extremely high among both sexes, males reported significantly higher than females (male 93.0% vs. female 82.6%, OR = 0.36, 95% CI: 0.18–0.67, *p* = 0.002). This means that females had 64% lower odds of heavy weight exposure than males. All other risk factors, specifically, salt exposure, working in wet clothes, lack of sanitary latrines, and absence of handwashing facilities, were found to be reported across both sexes at consistently higher rates with no statistically significant differences (see Fig 3).

**Table 4.**
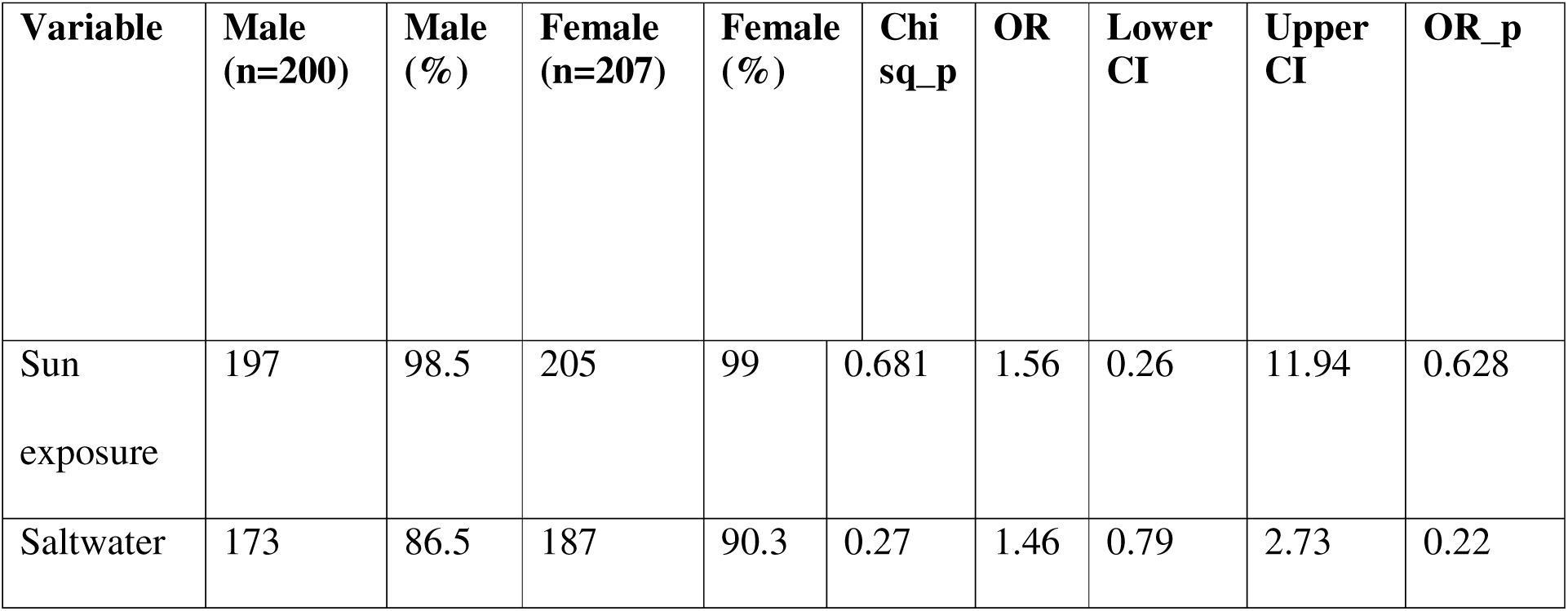
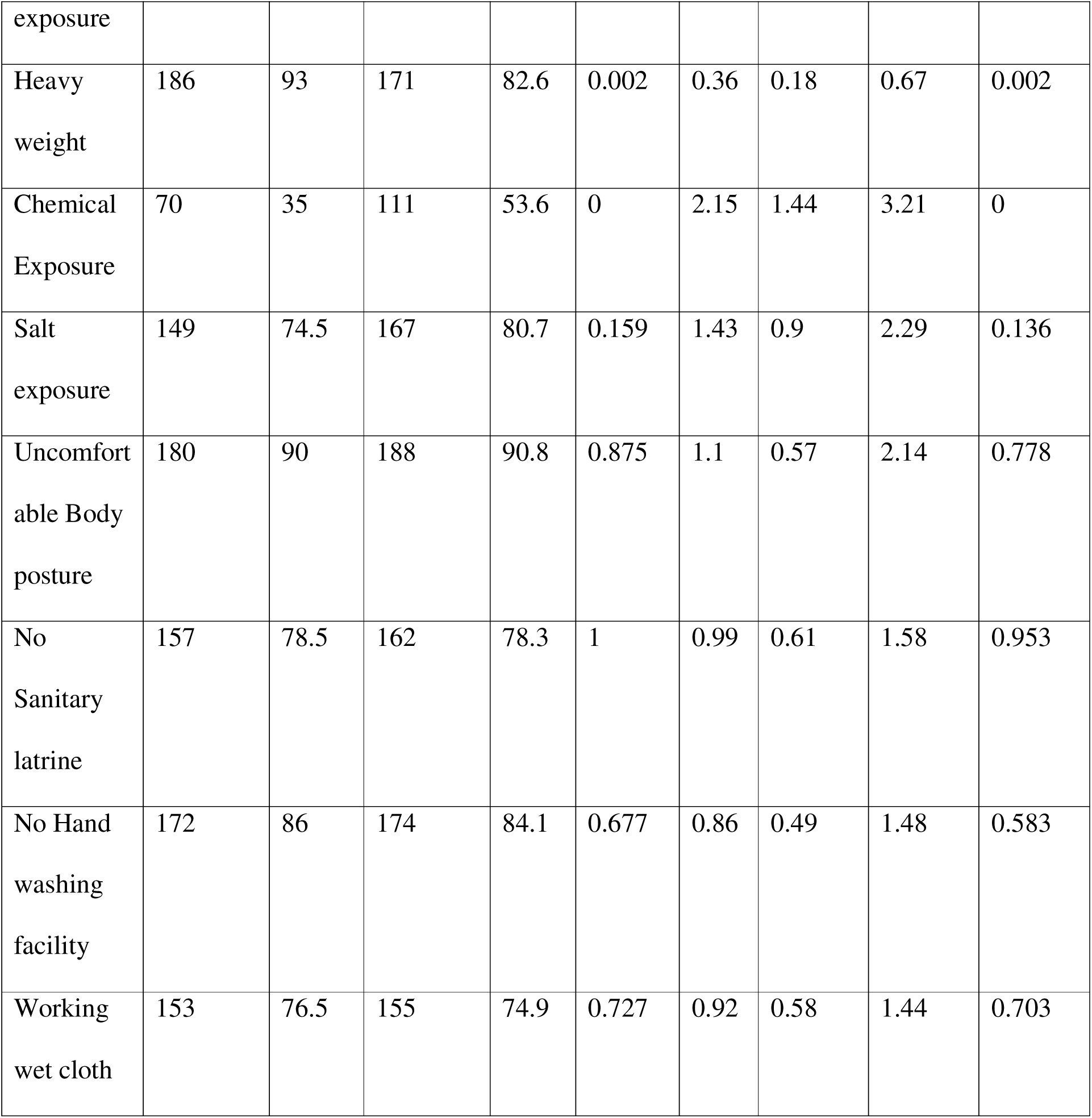
Prevalence of occupational hazards among men and women

**Figure 3.** Occupational hazard prevalence among male and female fisherfolk. Radar chart illustrating the reported prevalence of sun exposure, chemical contact, heavy lifting, and other workplace hazards.

Table 5 lists the comparative prevalence of self-reported health conditions among men and women working in the fisheries sector. Although no significant difference was observed across men and women, we found a very high prevalence of skin diseases, musculoskeletal issues, vertigo, diarrhoea, scabies, tinea among both groups, indicating both groups have the comparable risk of those health conditions.

**Table 5.** Prevalence of self-reported health conditions among men and women

| Variable | Male (n) | Male (%) | Female | Female (%) | Chi sq_(p) | OR | Lower CI | Upper CI | OR (p) |
| --- | --- | --- | --- | --- | --- | --- | --- | --- | --- |
| Fish related injury | 179 | 89.5 | 181 | 87.4 | 0.547 | 0.82 | 0.44 | 1.5 | 0.516 |
| Fish sting bite | 147 | 73.5 | 120 | 58 | 0.002 | 0.5 | 0.33 | 0.75 | 0.001 |
| Accidental fall | 2 | 1 | 6 | 2.9 | 0.285 | 2.96 | 0.67 | 20.34 | 0.188 |
| Eye disease symptoms | 89 | 44.5 | 110 | 53.1 | 0.089 | 1.41 | 0.96 | 2.09 | 0.082 |
| Skin disease | 102 | 51 | 117 | 56.5 | 0.272 | 1.25 | 0.85 | 1.85 | 0.264 |
| Skin allergy | 79 | 39.5 | 87 | 42 | 0.623 | 1.11 | 0.75 | 1.65 | 0.604 |
| Skin ulcer wound | 15 | 7.5 | 22 | 10.6 | 0.291 | 1.47 | 0.74 | 2.97 | 0.275 |
| Headache | 124 | 62 | 146 | 70.5 | 0.085 | 1.47 | 0.97 | 2.22 | 0.069 |
| Sunburn | 116 | 58 | 123 | 59.4 | 0.844 | 1.06 | 0.71 | 1.57 | 0.771 |
| Earache | 21 | 10.5 | 10 | 4.8 | 0.036 | 0.43 | 0.19 | 0.92 | 0.035 |
| Reduced hearing | 35 | 17.5 | 27 | 13 | 0.21 | 0.71 | 0.41 | 1.22 | 0.212 |
| Vertigo | 61 | 30.5 | 56 | 27.1 | 0.444 | 0.85 | 0.55 | 1.3 | 0.443 |
| Musculoskeletal issue | 136 | 68 | 143 | 69.1 | 0.838 | 1.05 | 0.69 | 1.6 | 0.814 |
| Fracture | 9 | 4.5 | 10 | 4.8 | 1 | 1.08 | 0.43 | 2.77 | 0.874 |
| Diarrhoea | 129 | 64.5 | 132 | 63.8 | 0.927 | 0.97 | 0.65 | 1.45 | 0.878 |
| Scabies | 102 | 51 | 108 | 52.2 | 0.84 | 1.05 | 0.71 | 1.55 | 0.813 |
| Tineasis | 105 | 52.5 | 106 | 51.2 | 0.849 | 0.95 | 0.64 | 1.4 | 0.794 |
| Chronic cough | 62 | 31 | 53 | 25.6 | 0.268 | 0.77 | 0.5 | 1.18 | 0.227 |
| Motion sickness | 108 | 54 | 81 | 39.1 | 0.003 | 0.55 | 0.37 | 0.81 | 0.003 |
| Reaction to smell | 180 | 90 | 186 | 89.9 | 1 | 0.98 | 0.51 | 1.88 | 0.961 |

Three specific conditions out of twenty health conditions were found to be significantly higher among male workers in the fisheries, these are – fish stings or bites, motion sickness and earache (see Fig 4). For fish stings or bites, the odds of females reporting fish bites or stings were half those of males (females 58.0% vs males 73.5%, OR = 0.50, 95% CI: 0.33– 0.75, *p* = 0.002). Motion sickness significantly has high prevalence among males (54.0%) in comparison to females (39.1%) (OR = 0.55, 95% CI: 0.37–0.81, *p* = 0.003). Although low prevalence, the odds of the females reporting earaches lower than males (10.5% vs. 4.8%, OR = 0.43, 95% CI: 0.19–0.92, *p* = 0.036).

**Figure 4.** Self-reported health conditions among male and female fisherfolk. Bar chart comparing prevalence of key health conditions, including skin disease, diarrhoea, motion sickness, and musculoskeletal pain.

### Focus Group Discussion Findings

The focus group discussion has yielded profound insights on the occupational hazards and risks encountered by both gender in the fishing communities. More severe injuries are commonly reported by men, resulting from slippery boat decks and accidents involving sharp tools. As one fisherman reported, “One in my boat slipped away from the boat and fell in the sea. His hand and leg were chopped into pieces by the blade of the boat.” Fish stings or bites or related injuries were also reported as a common risk that prevailed among men. This is attributed to direct handling of live fish at sea, as reported by one participant, “I catch fish from the sea. When we remove the fish from the net, many of them bite or sting.” Men also get exposed to dangerous species such as guijja, boiragi, and shaplapata. As per the fishermen, some stings lead to swelling, delirium, or prolonged illness.

Fishermen also reported engine smoke and prolonged exposure to loud noise as health hazards in their occupation, which often cause irritation in the eyes, persistent cough, and hearing loss. “When the engine works, we can’t even hear someone speaking beside us. One person became deaf.” reported by a fisherman. They also frequently reported low back pain, especially, those operating the boat or managing the nets. Pain worsens during rest days, as one reported, “When we are off duty, the back pain becomes unbearable.” Fishermen need to work for a prolonged period under the open sky in the deep sea or river for fishing, exposing them to the sun, which often leads to sunburn, dizziness, and dehydration.

During focus group discussions, women explained that due to their engagement in fish handling and dry fish processing, they often become exposed to fish slime, fermented shrimp, pesticides, salt, and chilli powder, which cause itching, allergic reactions, peeling skin, and even bleeding wounds. A woman involved in fish processing reported, “Hands burn, crack, and even bleed. I couldn’t touch rice properly for three months. I had to take medicines.” During fish processing, women also experience fish stings and cuts from species like *Churi*, *Tengra*, *Loitta*, and *Guijja*. In some cases, these lead to infection, pus formation, and fever. “Sometimes fish bone or teeth remain inside the skin it swells up and causes a lot of pain. We cut the hand if it gets worse.” Those engaged in Nappi processing reported that their exposure to gas released from Nappi often causes eye burning, blurred vision, and breathing difficulty. “Nappi releases pungent vapors. My eyes feel dry and itchy. Older women now have vision problems and asthma.” Women also reported musculoskeletal complaints, like knee swelling, lower back pain, and shoulder strain, which they think result from long working hours and working in squatting or bent positions. “We sit in one position for hours. The pain lasts even when we get home.”

Some women reported that poor access to toilets forced them to hold urine for a long time, causing urinary tract infections. “We hold urine for a long time while working. It causes burning when urinating. The toilet is shared, and there’s no soap.” They also invariably reported unavailability of menstrual hygiene kits in the workplace. Nursing women do not find a good and private place for breastfeeding their children, “there’s no place to breastfeed. We leave the baby nearby while we work on Nappi.” There were concerns about the lack of maternity leave. They usually did not receive any paid leave for childbirth. They were expected to return to work as soon as possible after childbirth.

Other gender-specific hazards for fishermen, especially those who fish in the deep sea, including drowning and falling overboard, cyclones and storms causing fear and danger, and attacks from pirates. In big boats, divers need to dive deep to untangle nets, putting them at great risk of strangulation in the nets, as the fishermen explained, “In big boats, the person who attempts to dive in case of a strangled net gets trapped and has died several times.”

Table 6 contains the work circumstances indicated by participants that resulted in fatalities among both men and women. Overall, the risk of fatality for men are reported to be significantly higher than for women.

**Table 6.** Cause of fatality among men and women

| Cause of Fatality Reported | Reported in Men | Reported in Women |
| --- | --- | --- |
| Drowning and propeller injury | Yes | Not mentioned |
| Strangulation in net | Yes | Not Mentioned |
| Diarrhoea / dehydration | Yes | Yes |
| Pirate attacks (Killing) | Yes | Not mentioned |
| Cyclone-related drowning/missing | Yes | Not mentioned |
| Cancer (linked to smoking) | Yes | Not mentioned |
| Fainting / Heatstroke | Yes | Yes |

### Personal Protective Equipment (PPE)

Table 7 shows the status of PPE use and first aid kit availability among men and women workers in fisheries. Overall, the use of PPE is low among both men and women, with 65% to 99% reporting not using the specific PPEs. Men, in significantly higher proportion, never wear masks in comparison to women (81.5% vs. 69.1%, p = 0.002). Although only a small fraction of men reported wearing gumboots (combined rarely and sometimes), it is significantly higher among men compared to women (11.00% vs. 5.3%, p = 0.004). Nearly all fishermen never used sunscreen, while a small fraction of women, 5.8%, reported at least rare use (p = 0.023). Most of both groups reported not using hand gloves, raincoats, and sunglasses, with no statistically significant differences by sex. While life jackets and life buoys are exclusively applicable to men, most of the fishermen did not use life jackets (59.0%) and life buoys (42.0%). A higher proportion of men have a first aid kit in their workplace (24.0%) compared to women (6.3%).

**Table 7:**
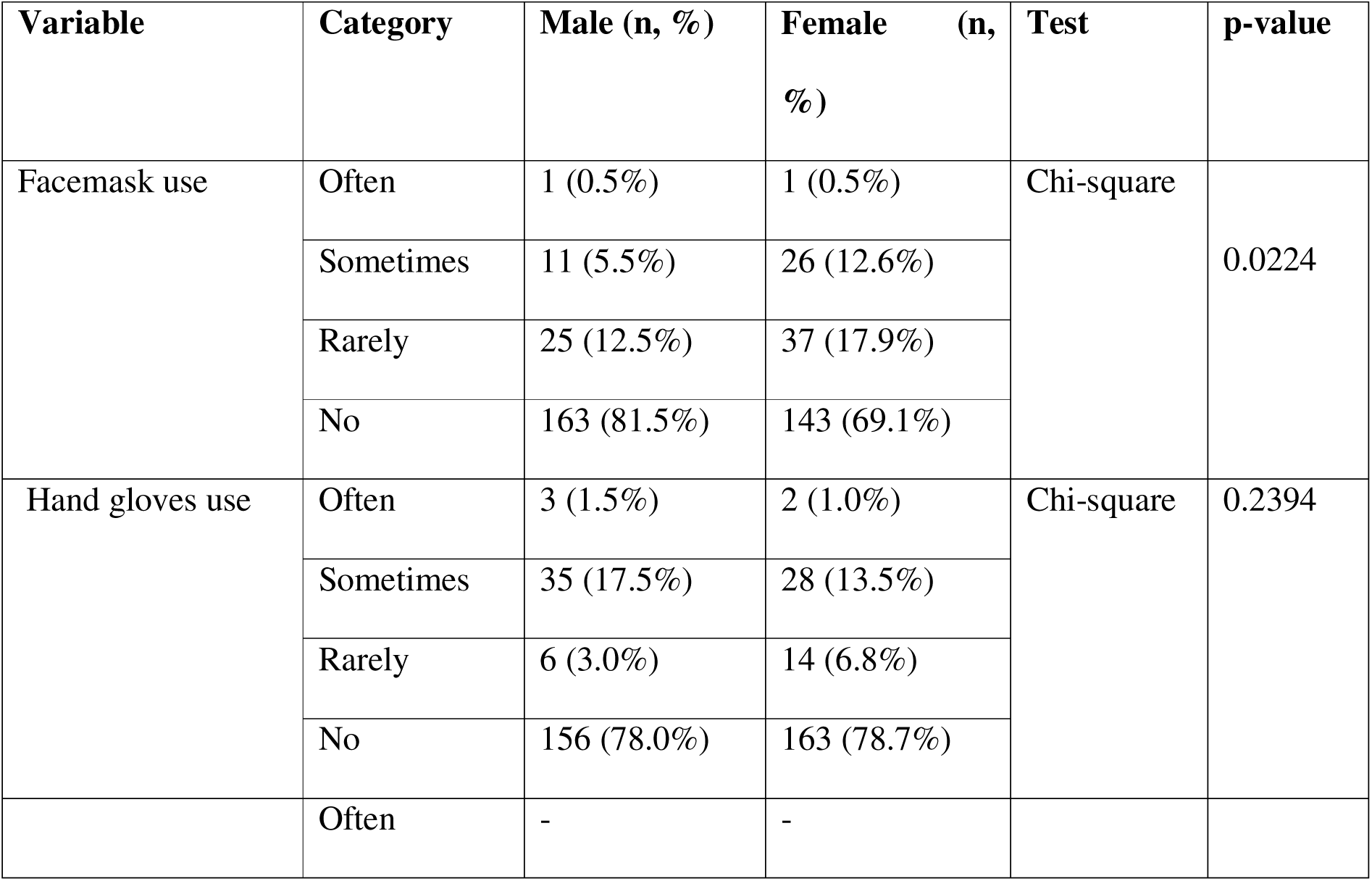
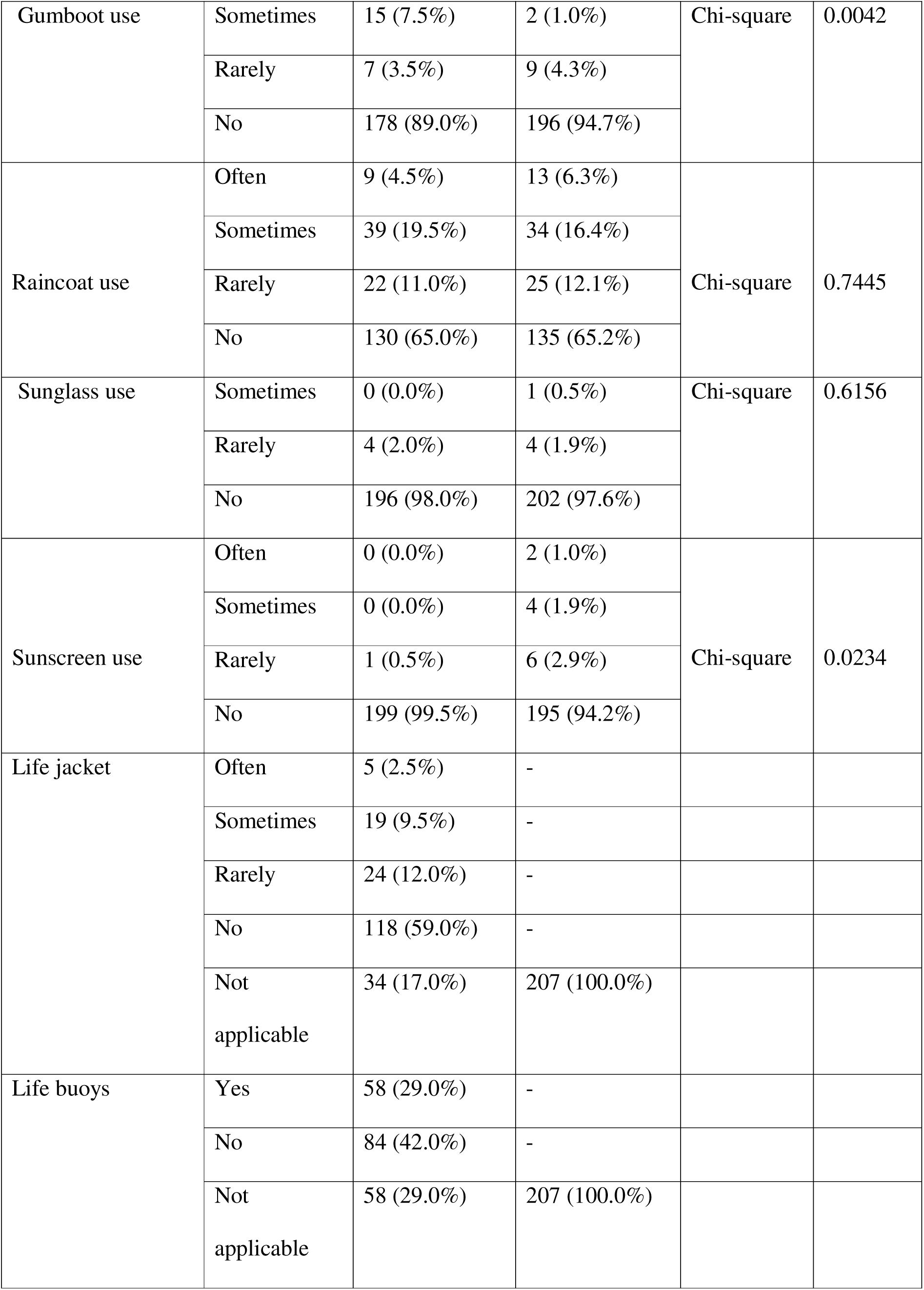
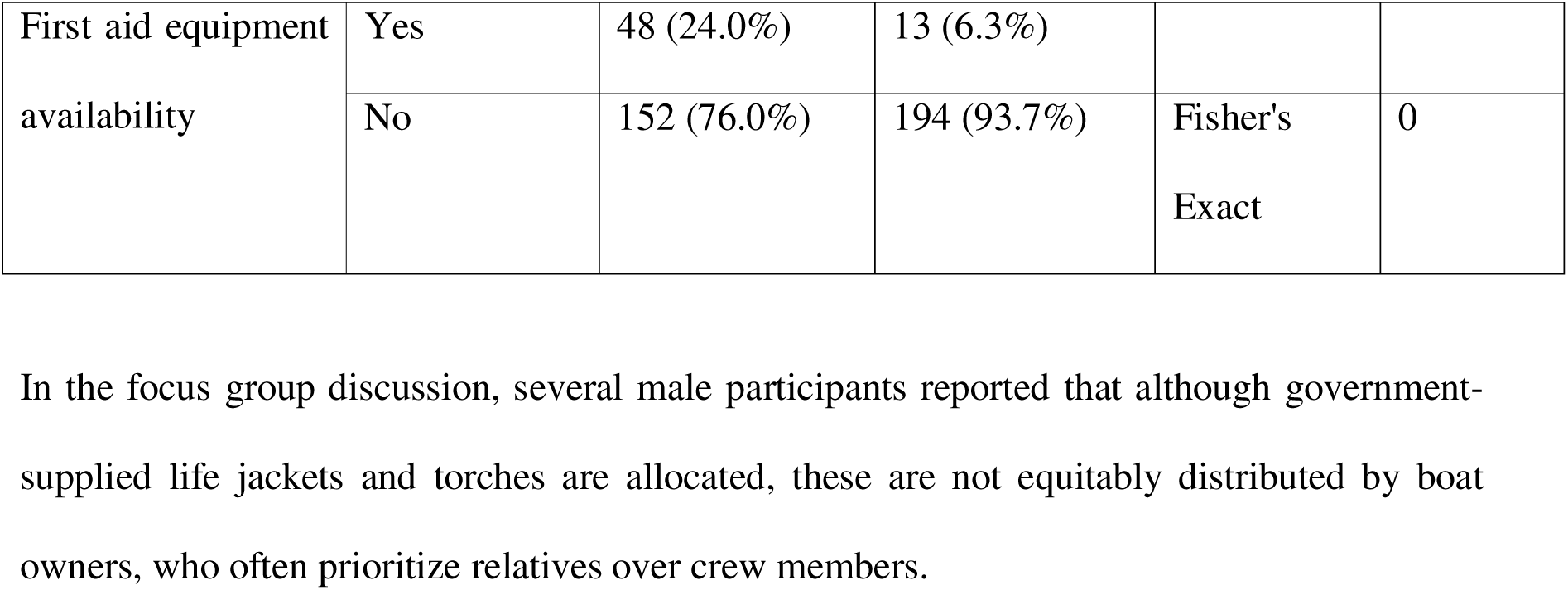
PPE use and first aid kit availability among men and women workers

In the focus group discussion, several male participants reported that although government-supplied life jackets and torches are allocated, these are not equitably distributed by boat owners, who often prioritize relatives over crew members.

## DISCUSSION

This is first study in Bangladesh and among few studies in developing countries that made a gendered comparison of occupational hazards and risks among the men and women workers in the fisheries sector. Overall, the study found a very high level of occupational hazards and risks prevalent among both genders, however, some hazards and risks are affecting men and women at different scale and impact. The nature and consequences of exposure to hazards and experience of disease or illness risks are found highly gendered. While fishermen experience more physical hazards and life-threatening risks at sea, female workers in fisheries experience prolonged chemical exposure and negligence in maternal and reproductive health.

We have found a high rate of illiteracy among both male and female participants, which could be resulted from as well result in poverty among the fisherfolk communities in Bangladesh. The high rate of illiteracy could also force men and women to work in hazardous working environments as well as result in their lower awareness of their occupational rights, safety measures and access to healthcare services, further exacerbating their vulnerability to occupational health risks (13, 22).

We have found a gendered segregation of occupational engagement of men and women in fishing communities. Men are exclusively engaged in direct riverine and offshore fishing. This role is socially constructed for men, since fishing requires physically demanding labour and highly associated with life-risks and culturally, masculinity is regarded as strength and bravery to accept this challenge(23, 24). Conversely, the study found women’s primary engagement in fish processing and coast-basd activities, and traditionally, an addition to maintaining their household responsibilities and caregiving roles. This is often associated with lower payment in comparison to men and unavailability of sick or maternity benefits. This finding aligns the previous studies in Africa, Asia-pacific and India as well as opinion of Food and Agriculture Organization that engagement of women in fisheries in considered as an extension of domestic work, and is undervalued from economic perspective (24–26).

The study revealed an extremely high prevalence of occupational hazards among both male and female fisheries workers. Workers of both genders have an alarmingly high rate of exposure to sun, salt, and saltwater without any statistical difference, meaning that both groups are equally exposed to the hazards. While the prevalence of exposure is almost the same, the setting and causes of exposure could be different. For instance, females are exposed to the sun due to their long hours of working under the open sky for dry fish processing, while men could be predominantly exposed due to their continuous working at boats in the sea. Such an extremely high prevalence of sun exposure among men and women in the fisheries may result in skin damage, sunburn, dehydration, heat exhaustion, and increased risk of skin cancer(27). This is also evident in our research as nearly sixty per cent of participants from both groups reported having sunburn as well as nearly or more than fifty per centage of symptoms of eye diseases and skin diseases without significant differences in prevalence. The findings are consistent with researches in India and Kenya (14,22), where high prevalence of sun exposure and resulting sun burn and ocular diseases were reported among fishers. Similarly, chronic contact with salt and salt water in day-to-day activities of men and women in the fisher folk could also be the contributing factors of high prevalence of skin, ocular and ear diseases found among men and women. While we acknowledge further research is warranted to establish the association of these hazards to the resulting disease risks, the researchers will try to address this in their upcoming third study of this umbrella research initiative.

We found that the women workers in fisheries are disproportionately affected by chemical exposure, including pesticides. This is explainable as women are more engaged in coast-based dry-fish processing activities. These findings align with previous studies from Bangladesh, India, Nigeria and Kenya indicating that women in fisheries are more exposed to hazardous chemicals due to their engagement in post-harvest tasks(9,28,29).Previous studies reported that occupational exposure to pesticides can be associated with chronic respiratory illnesses, including lung cancer (30). We have also seen a high prevalence of chronic cough among the men and women in fisheries. Since the scope of this paper to assess the gender-aspect of occupational hazards and risks, we will explore the impact of chemical hazards among the workers in fisheries in our upcoming study in this umbrella research initiative.

The study also revealed that a high rate of musculoskeletal issues exists among both men and women in the fisherfolk. While lifting heavy weights is common in both gender groups, it is still significantly higher among men (93% vs 83%, p value = 0.002). Among women, the musculoskeletal health issues could be linked to the prolonged working in awkward posture as identified in our previous study that they have to sit on their feet without any support to the hip or back. Also, this could be related to bearing heavy weight, like fish and fish products, in their processing tasks (13, 31). The dual burden of domestic responsibilities and economic laborious works can further exacerbate their physical strain (5,13). Conversely, in fishermen this could primarily be linked to lifting heavy weights, operating boats and hauling nets as revealed in our focus group discussions. This finding is consistent with the studies in other similar settings, like Greece, Kenya, India and Bangladesh(22,32), where also similar musculoskeletal risks among men and relevant hazards were also identified.

We have seen a very high prevalence of communicable diseases among both gender groups. For instance, more than sixty percent of men and women suffered from diarrhoea in last 12 months. This could potentially be linked to the fact that more than three fourth of the participants from both men and women groups did not have sanitary latrines, and nearly 85% did not have handwashing facilities at their workplace. We have also found more than half of the men and women participants reported scabies and tineasis in last 12 months. This could be potentiality linked to the fact that both groups are equally exposed to salt water and need prolong working in wet cloth (14,33). Lack of hygiene, for instance, inability of changing clothes and daily bath due may further exacerbate the risk of these communicable diseases.

The study found some unique challenges experienced by women workers in fisheries sector in relation to the reproductive health and maternity rights for women. Holding urine for extended periods due to poor access to safe and private toilet facilities lead the women to urinary tract infections. Absence of menstrual hygiene kits at workplace, private space for breastfeeding and absence of maternity leave as found in this study may affect the occupational health, well-being and dignity of the women(34–36). Policy reforms is critical in fisheries sector to address these structural inequities to create gender-sensitive sanitation facilities, workplace breastfeeding spaces, menstrual hygiene kit supplies, and provision of maternity benefits.

While we have seen very high incidence of fish related injuries among both men and women, men are more susceptible to experience injuries from fish sting or bite (73.5% vs 58.0%, p value 0.001). This can be explained from the fact that male is more engaged in handling live fish during fishing, giving them the higher risk of getting fish bite or sting (21). On the other hand, incidence of any type of injury related to fisheries appears similar among both groups because women are in higher risk of injury from fish processing instruments (like sharp instruments) and exposure to contact to fishes causing allergic reaction during fish processing, like removing from nets, grading and sorting (13). Men is also found having higher risk of experiencing earache in comparison to women (10.5% vs 4.8%, p value 0.035), which could potentially be attributed to their exposure to the noise from engines in the boat. The study also found that men are more exposed to life-threatening risks, such as drowning and falling overboard, especially during cyclones and storms, pirate attacks and entrapment in nets. This sometime leads them to fatalities as revealed during the focus group discussions. This gendered risk pattern stems from societal norms that position men predominantly in offshore and deep-sea fishing in offshore and deep-sea fishing demanding physical effort and expecting the masculine character of risk taking. The similar finding has also been reported in another study among fishermen in Bagerhat, Bangladesh (9) as well as studies in coastal Ghana and the Philippines, where men face higher fatality rates due to extreme weather events and occupational hazards at sea (37, 38). Promoting early warning and alert system, developing and supplying safety equipment, designing safer vessels, educating fishers on safety and personal protection and organizing capacity building initiatives are warranted to prevent such live-threatening conditions and fatalities (39,40).

Many of the hazards could be effectively eliminated by use of the PPEs , specially, protective gloves, eye goggles, boots and masks, preventing the occurrence of diseases and illnesses (22, 41). However, our study found that despite high prevalence of occupational hazards among both men and women workers in fisheries, a high proportion of men and women are not using PPEs. The finding is consistent with other similar settings in low and middle-income countries, specially, India, Kenya, Gambia and Thailand(19, 42). In women, this is related to lack of supply of the PPEs and the fact that commonly available PPEs (e.g. heavy-duty gloves) may not be suitable use for their fine work during fish processing(13). For fishermen, although there is allocation of PPE from the government, the issue of low use can be related to lack of compliance by the boat owner for distribution and by the fishermen to use. Therefore, to prevent occupational risks among the men and women working in fisheries, the strong monitoring system should be in place, customized PPEs should be innovated and tailored to the needs of the users, and all stakeholders in fisheries should be sensitized on PPE and safety measures.

## LIMITATIONS

The research identified the hazards and health risks based on the experiences shared by the participants, not through clinical examination or validation. Authors tried to mitigate the response bias by triangulation of qualitative and quantitative data and proper training of the enumerators. The clinical correlation between identified hazards and specific health outcomes was beyond the scope of this study and will be addressed in next research under this umbrella research initiative.

## CONCLUSION

This study revealed a high burden of occupational hazards and risks among both male and female workers in the fisheries sector in Bangladesh from a gender lens. The study revealed that a large number of hazards and health risks among both male and female workers requiring urgent attention for both groups. Among the unique challenges, men are predominantly exposed to physical and life-threatening risks at sea, while women face prolonged chemical exposure, and violations of reproductive health rights in processing tasks. A high illiteracy coupled with limited use of personal protective equipment excerbates the consequences of the hazards and risks. There is an urgent need for gender-sensitive occupational safety and health strategy and action plan for the fisheries sector highlighting prevention of hazards, provision of employment benefits, innovation and supply of contextualized personal protection equipment, recognizing women’s role in fisheries and promoting their maternal and reproductive health and reducing the life-threatening risks of fishermen. Such gender sensitive strategy is crucial to attain the health, wellbeing, dignity, productivity, and protection of the men and women in fisherfolk communities in Bangladesh.

## FUNDING

This study received no specific grant from any funding agency in the public, commercial or not-for-profit sectors.

## COMPETING INTEREST

The authors declare no competing interests. This study received no external funding.

## Data Availability

All data generated or analyzed during this study are available as follows:
Quantitative survey data: Publicly accessible on Zenodo at https://doi.org/10.5281/zenodo.15689935
Qualitative data (FGDs): Due to confidentiality and participant privacy concerns, the full transcripts are not publicly shared but are available from the corresponding author upon reasonable request.

## SUPPORTING INFORMATION

**S1 File.** Structured questionnaire for male participants. This questionnaire was used during the quantitative survey of male fisherfolk in Chattogram and Cox’s Bazar.

**S2 File.** FGD guide. The semi-structured questionnaire used for conducting focus group discussions with male and female fisherfolk.

## Notes

### Competing Interest Statement

The authors have declared no competing interest.

### Funding Statement

As the corresponding author and all the co-authors are based in Bangladesh (a Group A country per Research4Life classification), we request a full waiver of the Article Processing Charge (APC). No grant or institutional funding is available to cover publication costs. We look forward to the opportunity to contribute to the important body of work published in PLOS ONE.

### Author Declarations

Ethics committee/IRB of North South University Institutional Review Board (NSU IRB), Dhaka, Bangladesh gave ethical approval for this work. The approved protocol number is 2023/OR-NSU/IRB/0810.

